# Impact of COVID-19 pandemic on lung cancer treatment scheduling

**DOI:** 10.1101/2020.06.09.20126995

**Authors:** Kohei Fujita, Takanori Ito, Zentaro Saito, Osamu Kanai, Koichi Nakatani, Tadashi Mio

**Author notes:** **Corresponding author** Kohei Fujita, MD, PhD, Division of Respiratory Medicine, Center for Respiratory Diseases, National Hospital Organization, Kyoto Medical Center, Address: 1-1, Fukakusa-Mukaihata, Fushimi-Ku, Kyoto, Japan.

## Abstract

**Objective:** Current pandemic of coronavirus disease 2019 (COVID-19) is associated with a heavy burden on the mental and physical health of patients, regional healthcare resources, and global economic activity. Many patients with lung cancer are thought to be affected by this situation. Therefore, we aimed to evaluate the impact of COVID-19 pandemic on lung cancer treatment scheduling.

**Study design:** We retrospectively reviewed the medical records of lung cancer patients who were undergoing anti-cancer treatment at the National Hospital Organization Kyoto Medical Center (600 beds) in Kyoto, Japan, between March 1, 2020 and May 31, 2020.

**Methods:** After the medical records were reviewed, the patients were assigned to one of two groups, depending on whether their lung cancer treatment schedule was delayed. We assessed the characteristics, types of histopathology and treatment, and the reason for the delay.

**Results:** A total 15 (9.1%) patients experienced the delay of lung cancer treatment during COVID-19 pandemic. Patients with treatment delay received significantly more ICIs monotherapy than patients without treatment delay (p=0.0057). On the contrary, no patients receiving molecular target agents experienced treatment delay during COVID-19 pandemic period (p=0.0027). The treatments of most of the patients were delayed per their request.

**Conclusion:** We revealed 9.1% lung cancer patients suffered anxiety and requested treatment delay during COVID-19 pandemic. Oncologists should keep in mind that patient with cancer have more anxiety than we expected under special occasions such as COVID-19 pandemic.

## Introduction

By the end of May 2020, coronavirus disease (COVID-19), which is caused by the severe acute respiratory syndrome coronavirus 2 (SARS-CoV-2), was reported to have affected more than 6,000,000 people globally (1, 2). COVID-19 is known to place a heavy burden on the mental and physical health of patients, regional healthcare resources, and global economic activity. In Japan, the government officially declared a state of emergency on April 7, 2020 (3). Daily coverage of the COVID-19 pandemic has been negatively affecting the mental health of patients. Considering that patients with cancer may suffer worse respiratory outcomes than those without (4, 5), this pandemic is presumed to affect the daily routines of cancer patients and oncologists. Therefore, in this study, we aimed to evaluate the impact of the COVID-19 pandemic on lung cancer treatment scheduling.

## Patients and methods

The period of the COVID-19 pandemic has been defined as the time between March 1, 2020 and May 31, 2020. We retrospectively reviewed the medical records of lung cancer patients who were undergoing anti-cancer treatment at the National Hospital Organization Kyoto Medical Center (600 beds) in Kyoto, Japan, between March 1, 2020 and May 31, 2020. The reviewing criteria were as follows: (A) pathologically confirmed lung cancer patients; (B) patients who received treatment with cytotoxic chemotherapy ± immune checkpoint inhibitors (ICIs), ICI monotherapy, or molecular target agents; and (C) patients who received treatment during the COVID-19 pandemic period. After the medical records were reviewed, the patients were assigned to one of two groups, depending on whether their lung cancer treatment schedule was delayed. We assessed the characteristics, types of histopathology and treatment, and the reason for the delay.

The medical records of 165 patients who met the inclusion criteria were reviewed. The characteristics of the patients are summarized in Table 1. The patients were predominantly male (mean age: 70.2 ± 9.2 years). Regarding lung cancer type, the highest proportion of patients had adenocarcinoma, followed by squamous cell carcinoma, small-cell lung cancer, and a type that was not otherwise specified. Of the 165 patients, 133 (80.6%) received their treatment at the outpatient clinic; 33 (20.0%), 76 (46.1%), and 56 (33.9%) patients had received cytotoxic chemotherapy ± ICIs, ICI monotherapy, and molecular target agents, respectively. This retrospective study was approved by the relevant institutional review board (approval number: 20-022).

**Table 1.** Characteristics of lung cancer patients undergoing treatment during the COVID-19 pandemic period

|  | All patients<br>n = 165 | Patients with treatment delay<br>n = 15 | Patients without treatment delay<br>n = 150 | p-value |
| --- | --- | --- | --- | --- |
| Age | 70.2 ± 9.2 | 72.5 ± 9.2 | 70.0 ± 9.2 | 0.28 |
| Sex (female) | 62 (37.6) | 3 (20.0) | 59 (39.3) | 0.17 |
| Outpatient treatment | 136 (82.4) | 11 (73.3) | 125 (83.3) | 0.30 |
| Histopathology |  |  |  |  |
| Adenocarcinoma | 99 (60.0) | 6 (40.0) | 93 (62.0) | 0.11 |
| Squamous cell carcinoma | 32 (19.4) | 5 (33.3) | 27 (18.0) | 0.17 |
| Small cell lung cancer | 18 (10.9) | 1 (6.7) | 17 (11.3) | >0.99 |
| Not otherwise specified | 15 (9.1) | 3 (20.0) | 12 (8.0) | 0.14 |
| Pleomorphic carcinoma | 1 (0.61) | 0 (0.0) | 1 (0.67) | >0.99 |
| Type of treatment |  |  |  |  |
| Cytotoxic chemotherapy ± ICIs | 33 (20.0) | 3 (20.0) | 30 (20.0) | >0.99 |
| ICI monotherapy | 76 (46.1) | 12 (80.0) | 64 (42.7) | 0.0057 |
| Molecular target agents | 56 (33.9) | 0 (0.0) | 56 (37.3) | 0.0027 |
| Reason for treatment delay |  |  |  |  |
| Doctor request | 2 (1.2) | 2 (13.3) | - | NE |
| Patient request | 12 (7.3) | 12 (80.0) | - | NE |
| Family request | 1 (0.61) | 1 (6.7) | - | NE |

Data are shown as number (%) or mean ± SD.
Abbreviations: ICI = immune checkpoint inhibitor, NE = not evaluated.

### Statistical analyses

A chi-square test or Fisher’s exact test was applied to compare the categorical variables. Alternatively, the Mann-Whitney U-test was applied for the continuous variables. The condition for statistical significance was defined as a p value below 0.05. Statistical analyses were performed using SPSS v26.0 (IBM SPSS, Inc., Chicago, IL, USA).

## Results

We reviewed the medical records of 165 patients who had undergone lung cancer treatment during the study period. The lung cancer treatments of a total of 15 patients (9.1%) were delayed during the COVID-19 pandemic. Patients with delayed treatment received significantly more ICI monotherapy than patients without delayed treatment (p = 0.0057). By contrast, no patients who received molecular target agents experienced a treatment delay during the COVID-19 pandemic period (p = 0.0027). Figure 1 shows the proportion of patients whose treatment was delayed according to the type of treatment. Of the 76 patients who received ICI monotherapy, 12 patients (15.8%) experienced treatment delay. However, the treatments of most of the patients were delayed per their request. Alternatively, the doctors of two patients recommended delayed treatment, and one patient was advised by their family to delay treatment.

**Figure 1.**
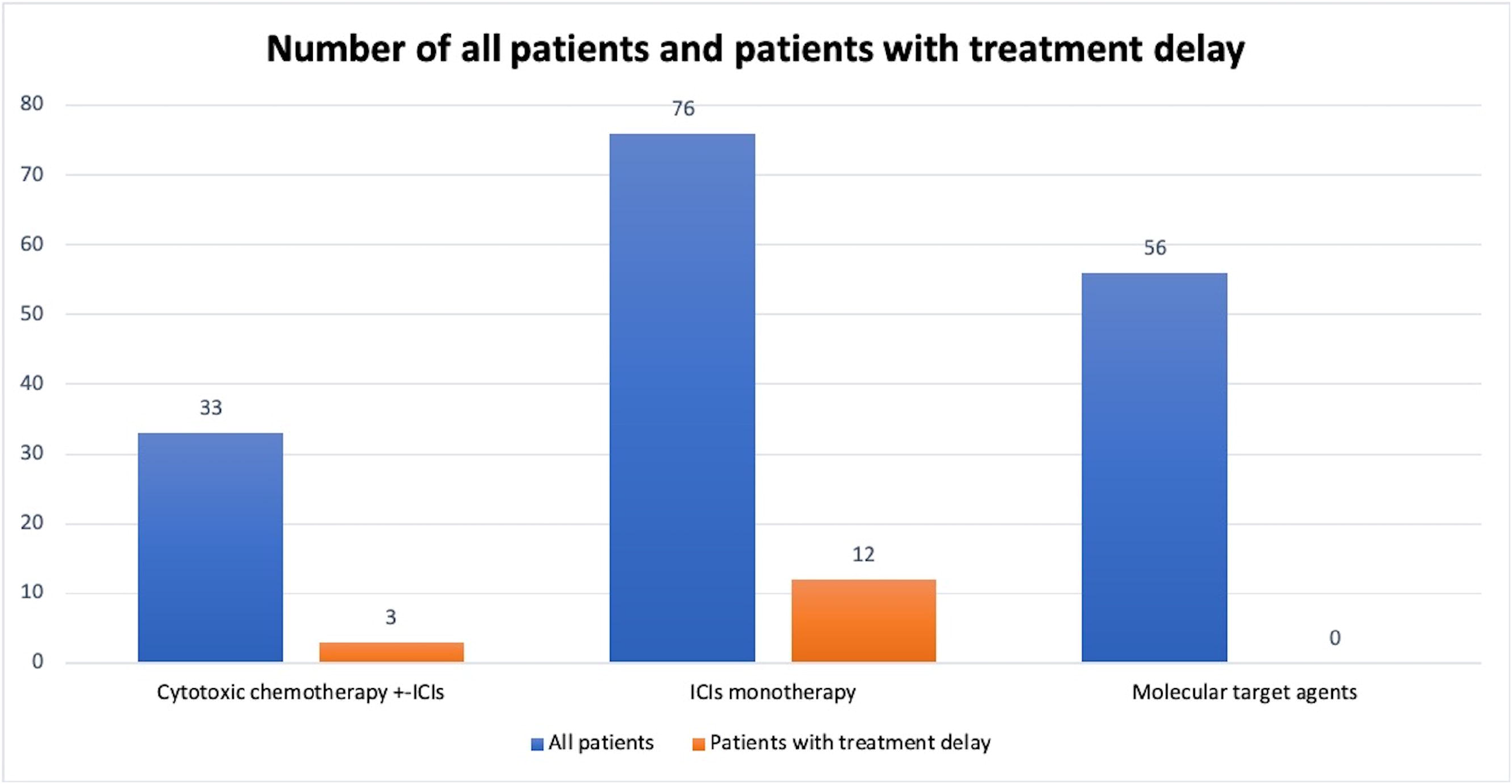
Proportion of patients who delayed treatment according to treatment type. The blue bar indicates the total number patients who received each type of treatment. The red bar indicates patients associated with delayed treatment.

## Discussion

Here, we revealed that the lung cancer treatment schedules of 9.1% of lung cancer patients were altered during the COVID-19 pandemic. In most cases, treatment delay was requested by the patient, suggesting that lung cancer patients had more COVID-19-related anxiety than expected. Several studies have indicated that, compared to individuals with no history of cancer, cancer patients have a higher risk of death if they are infected with SARS-CoV-2 and develop COVID-19 (4, 5). In addition to host traits, hospital admission and recurrent hospital visits are also potential risk factors for SARS-CoV-2 infection (6). Because the media has widely disseminated this information, many cancer patients are believed to experience anxiety regarding COVID-19. Although several countries have advocated suggestions for cancer treatment during the COVID-19 pandemic advocated from several countries (7, 8), a consensus has not been reached; thus, there is no generalized standard for cancer treatment during this pandemic.

In our study, among the patients receiving one of three types of treatment agents, a higher proportion of patients who received ICI monotherapy delayed their treatment.

Recently, an expert advocated weight-based dosing of pembrolizumab (9). According to their recommendation, patients should receive a 400-mg dose every 6 weeks instead of a 200-mg dose every 3 weeks. This can reduce the number hospital visits and lower the risk of infection. Various science-based plans and ideas such as this will be required during a pandemic period.

This study had several limitations. First, this study was retrospectively conducted in a single center, which caused selection bias. Specifically, the proportion of delayed treatment cases is dependent on the severity of the epidemic in an area. Kyoto has been deemed to be a mild epidemic area. Thus, it is expected that areas where the severity of the COVID-19 pandemic is higher will be associated with more patient requests for treatment delay. Second, because the COVID-19 pandemic is unprecedented and ongoing, the study period may have been underestimated. Lastly, the effects of treatment delay on the prognosis of lung cancer are unknown. Nevertheless, our study provides insight into the COVID-19 pandemic impact on cancer treatment.

In conclusion, we revealed that 9.1% of lung cancer patients suffered anxiety and requested treatment delay during the COVID-19 pandemic. Therefore, oncologists should keep in mind that cancer patients tend to have more anxiety than is expected under special circumstances such as the COVID-19 pandemic.

## Data Availability

After approval from our institutional review board, this study data can be shared with qualifying researchers who submit a proposal with a valuable research question.

## Author contributions

KF designed and conducted this study. KF, TI, and ZS reviewed patient medical records. OK and TM reviewed and checked the collected data. KF drafted and revised manuscript. All authors approved this manuscript.

## Acknowledgments

We would like to thank Drs. Yuki Yamamoto, Misato Okamura, and Akihiro Yasoda for their helpful feedback on this work.

